# Effect of an online digital educational intervention on knowledge and perception of nursing students in Nigeria towards research: a pre- and post-test study

**DOI:** 10.1101/2025.10.26.25338852

**Authors:** Bose C. Ogunlowo, Oluwadamilare Akingbade, Emmanuel O. Adesuyi, Yetunde O. Tola, Khadijah Jimoh, Jessica O. Esangbedo, Esther Adeshina, Rebecca Oyedele, Damilare Aduroja, Tomiwa Olusoji, Damilola R. Okunola, Ruth O. Ololade, Victoria Faremi, Damilola A. Adebiyi, Iyabo Y. Ademuyiwa

## Abstract

Nursing research is a scientific process crucial for evidence-based practice. Learning about the research process empowers nursing students to comprehend research, actively engage in research endeavours, and integrate findings into clinical practice. Although evidence suggests that educational interventions might improve the research competencies of nursing students, little is known regarding the impact of such interventions on this population, which prompted this study, which aimed to ascertain the effects of an online digital educational intervention on the knowledge and perception of nursing students in Nigeria towards research.

The study utilized a pre-test and post-test approach. Nursing students between the second and final year of their studies were included. One hundred fifty nursing students received the educational intervention on June 17, 2023, while One Hundred thirty-five completed the pre- and post-test through an online questionnaire. Descriptive statistics were used to summarise the main features of the data, while hypotheses were tested using the Mann-Whitney U test. The majority of the respondents were females (75.3%), between 20-29 years (84.0%), Yoruba (56.0%), from the South-western part of Nigeria (46.7%), in the third year of training (51.3%), attend a school of Nursing (57.3%), and private-owned institutions (48.0%). We discovered a significant difference between the research knowledge of Nigerian nursing students before and after the intervention (p = <.001), with a medium effect size (*d* = 0.55). Similarly, there was a significant difference between the research perception of Nigerian nursing students before and after the intervention, *p* = .012, with a medium effect size of (*d* = 0.24).

The online digital educational intervention significantly increased knowledge and positively changed the perception level of Nigerian nursing students toward research. This suggests that similar interventions could greatly benefit Nigerian nursing students in the future, enhancing their research competencies and ultimately improving patient care.

**Author summary:** Strong research skills are essential for improving patient care. However, many nursing students in Nigeria find research challenging and see it mainly as an academic requirement rather than a vital part of clinical practice. This inspired us to design and test an online educational program to make research more accessible and engaging for nursing students.

In this study, we worked with nursing students across Nigeria, guiding them through an online session on the research process, from choosing a topic to writing and publishing their work. Before and after the session, students completed questionnaires that measured their knowledge and perceptions of research. The results showed a clear improvement: students gained more understanding of research and developed a more positive attitude towards it.

These findings suggest that digital learning can help bridge knowledge gaps and prepare future nurses to apply research evidence in patient care. By integrating online learning into nursing education, we can empower students to see research not as a burden, but as a tool for transforming healthcare practice.

## Introduction

Clinical nursing practice benefits from new knowledge discovered, improved, and validated through the scientific process of nursing research [1]. Current research advances evidence-based practice in nursing, which emphasises comprehensive and high-quality patient-centered care, emphasizing the patient rather than conventional practices, peer recommendations, or personal convictions [2]. Through collecting, evaluating, and utilizing research findings, nurses can expand their knowledge and improve their experience in clinical practice [2].

Although studies have shown that Nigerian nurses have a good impression of research conduct and feel that research is helpful and promotes nursing care practices [3–8], unfortunately, few nurses are actively involved in research; meanwhile, most nurses in Nigeria who have participated in research either gather data, organize studies conducted by physicians and other healthcare professionals, or satisfy a degree requirement. [9–10,8,11–12]. Studies on research conduct and usage among Nigerian nurses indicated that a small percentage had performed research and applied the findings to clinical practice [9,11]. Moreover, the absence of a thorough national registry of nurse researchers and the studies they have undertaken makes it challenging to determine the number of nurses engaged in academic research in Nigeria [8]. The positive perception of nurses and their understanding of the value and significance of nursing research reflects that although Nigerian nurses do little nursing research, a large portion would like to do so if given the necessary training and nursing education [6]. Nursing education provides the skills and knowledge necessary to advance nursing and health care. It also lays the groundwork for nursing research [10].

The first step in arousing nurses’ interest in research is training them during undergraduate education [13]. Learning about research equips nursing students with an understanding of research, which enables them to value it, actively engage in research, and incorporate research findings into practice [13]. Nursing students should have a good understanding of the stages of research, understand and critique research reports, and be able to apply research findings in achieving evidence-based practice to enhance the quality of life for patients [14,13). Similarly, adequate knowledge and positive perceptions of nursing students toward research will promote evidence-based practice [15].

Online learning is a network-enabled transfer of information and skills that allows education to be delivered to a large number of students at the same time or at different times. It offers opportunities for the development of new teaching resources, interactive learning, self-paced learning, and convenient access [16]. The use of digital learning in nursing education accelerated during the coronavirus pandemic when remote forms of learning were required to keep students and educators safe [17]. Virtual education techniques are used to increase learners’ knowledge and skills by using appropriate technology and facilitating the learning process at any time and place. The use of technology in education is necessary to improve the knowledge and perception of Nurses, both in learning and practice [18, 19].

Digital learning offers synchronous communication between learners and teachers, and can be via audio calls and video conferences, enhancing the learning experience. However, despite their similarities to real classrooms, they have limitations such as technical issues and not fitting all learning styles. Despite these challenges, advancements in technology could address these issues, and online learning can improve skills and increase confidence [19].

Akingbade et al. [9] suggested that online educational interventions might improve the research competencies of Nigerian nurses and nursing students, little is known about the effect of such intervention in this population, which prompted this study.

This study aimed to assess the effects of an online digital educational intervention on the knowledge and perception of nursing students in Nigeria towards research. It is also expected to provide significant insight into the factors influencing students’ perception of research, which could be relevant in proffering solutions to most nursing students across nations who perceive research to be difficult, challenging, and almost impossible due to insufficient knowledge. This study will broaden the body of research knowledge and encourage nurse educators to explore various ways of teaching to make students understand research better.

## Materials and methods

### Study design

This study used a pre-test and post-test approach, administering an online structured questionnaire to evaluate how the educational intervention affected the knowledge and perception of nursing students in Nigeria towards research.

### Setting and sample

This study utilised a total population sampling of 135 nursing students who were part of the webinar attendees and intended to participate in the pre-and post-test.

The inclusion criteria were being a nursing student between the second and final year of study in any post-secondary institution accredited by the Nursing and Midwifery Council of Nigeria and willing to participate in the educational webinar of this study. The students involved were not accepted into an exclusive program at the point of data collection. Hence, they were not pressured/coerced to participate in the study. In contrast, students who did not meet the category above were not allowed to participate in the study.

### Intervention

#### Format and duration

An online educational intervention was conducted on June 17, 2023, with the theme “Research without tears: Demystifying the research process for nursing students.” The intervention was delivered via Zoom and lasted four hours. Each session consisted of 45 minutes of presentations, 15 minutes of question-and-answer time, and a 20-minute break between each session.

#### Strategy/Content

Each session was facilitated by registered nurses who had obtained a doctorate/in a doctorate program at the time of the webinar. The intervention comprises three major sessions/lines of focus: the overview of nursing research, research methodology, and manuscript writing and publication. The overview section of the research process focused on selecting a desirable research area and topic, sampling design process methods, literature review, challenges encountered in nursing research, and strategies to overcome them.

The methodology session detailed how to make good methodological choices by knowing the research paradigm, choosing a research approach and appropriate design, suitable sampling, data collection, and analytical techniques. Furthermore, the manuscript writing and publication session exposed the participants to various scholarly papers, journals, and selection criteria. Similarly, the participants were enlightened on how to detect predatory journals and how to handle possible rejections. The session also buttressed the importance of research publication and its dissemination to the target populations, stakeholders, and policymakers, which will, in turn, fast-track the decision-making process in clinical practice and patient care.

### Instrument for data collection

Data collection for this study was through a well-structured questionnaire in a close-ended form to assess the research study’s format, layout, and relevance. The validated instrument used for data collection in this study was adapted from the study of Wu et al. [20]. The adapted questionnaire consisted of three parts, comprising 36 items. Section A consists of questions assessing the sociodemographic data of the participants with six items. Section B consists of twenty-five (25) items that elaborate on the information regarding knowledge of the research process. In this section, fifteen of the twenty-five questions used an Agree/Disagree/Not Sure structure. A score of 1 was given for the correct answer to each of the 25 questions, while a score of 0 was given to other options. Scores of 14-25 were considered to have a high level of research knowledge, while 0-13 were considered to have a low level of research knowledge. Section C consists of questions that evaluated the perception of nursing students toward research with five items. Strongly disagree, agree, not sure, agree, and strongly agree are the options for these five questions. Strongly agreeing resulted in a score of “4” on positively framed questions, whereas strongly disagreeing resulted in a score of “1”. For negatively framed questions, the “strongly disagree’’ option was given a score of “4”, whereas the “strongly agree” option was given a score of “1”. For positively and negatively worded questions, 0 was assigned to Not sure. Scores of 8-20 were considered a good perception of research, while 0-7 were considered a poor perception.

A pilot study was conducted among 17 nursing students not part of the main study to ensure reliability. The data received were analyzed to ascertain the instrument’s consistency, and a value of 0.78 was found using Cronbach’s alpha reliability test, which is significant for the instrument’s reliability.

### Data collection/procedure

An online survey via Google Forms was used for data collection. Participants were invited to register to attend a webinar on nursing research. During their online registration, they were asked if they would like to participate in research related to the Webinar. Those who responded ‘Yes’ to that question received a unique link in their email to access the Google form, which contains the pre-test questions before the webinar begins. The landing page of the Google form included written research information and informed consent details, outlining participants’ confidential and refusal rights, which they responded to by clicking the ‘agree’ or ‘disagree’ button. Participants who agreed to continue with the study entered the survey platform, where they responded to the questionnaire. All pre-test data were completed before the commencement of the webinar. After the webinar, participants received another link to respond to the post-test questionnaire almost immediately. Responses were compiled and coded correctly in an Excel sheet that is password-protected and accessible only to the research team. Since this is an online data collection format, the research team encountered challenges related to the omission of questions. Hence, only fully completed questionnaires were compiled, while incomplete questionnaires were considered invalid.

### Method of data analysis

Data were analyzed using version 25 of the IBM Statistical Package for Social Sciences (SPSS) software. The research questions were described using descriptive statistics, which included frequency distribution, means, and standard deviations, and the data were displayed through tables and charts. The Mann-Whitney U Test was used to determine the inferential statistics by comparing the pre-test and post-test knowledge and perception scores before and after the educational intervention at a significant level of *P<* .05

### Ethical Consideration

The Lagos University Teaching Hospital Health Research Ethics Committee, Idi-Araba, Lagos State, granted ethical approval to conduct this study (ethical approval number ADM/DSCST/HREC/APP/6322). Informed consent was obtained from all participants before their involvement in the study. Participants were fully informed about the purpose of the research, the voluntary nature of their participation, and their right to confidentiality and refusal at any point in the study.

## Results

This study used a total population sampling of 135 nursing students who participated in the webinar and fully completed the pre-test and post-test questionnaire forms. The response rate was 100%. The sociodemographic characteristics of the respondents included their gender, age, ethnic group, geopolitical zone of residence, institution, and ownership of the institution. Table 1 shows the sociodemographic characteristics of 135 sampled respondents. The participants were mostly females (75.6%), between 20 and 29 years old (81.5%), Yoruba (53.3%), from the Southwest (46.7%), School of Nursing (55.6%), and attending private-owned institutions (48.1%).

**Table 1.**
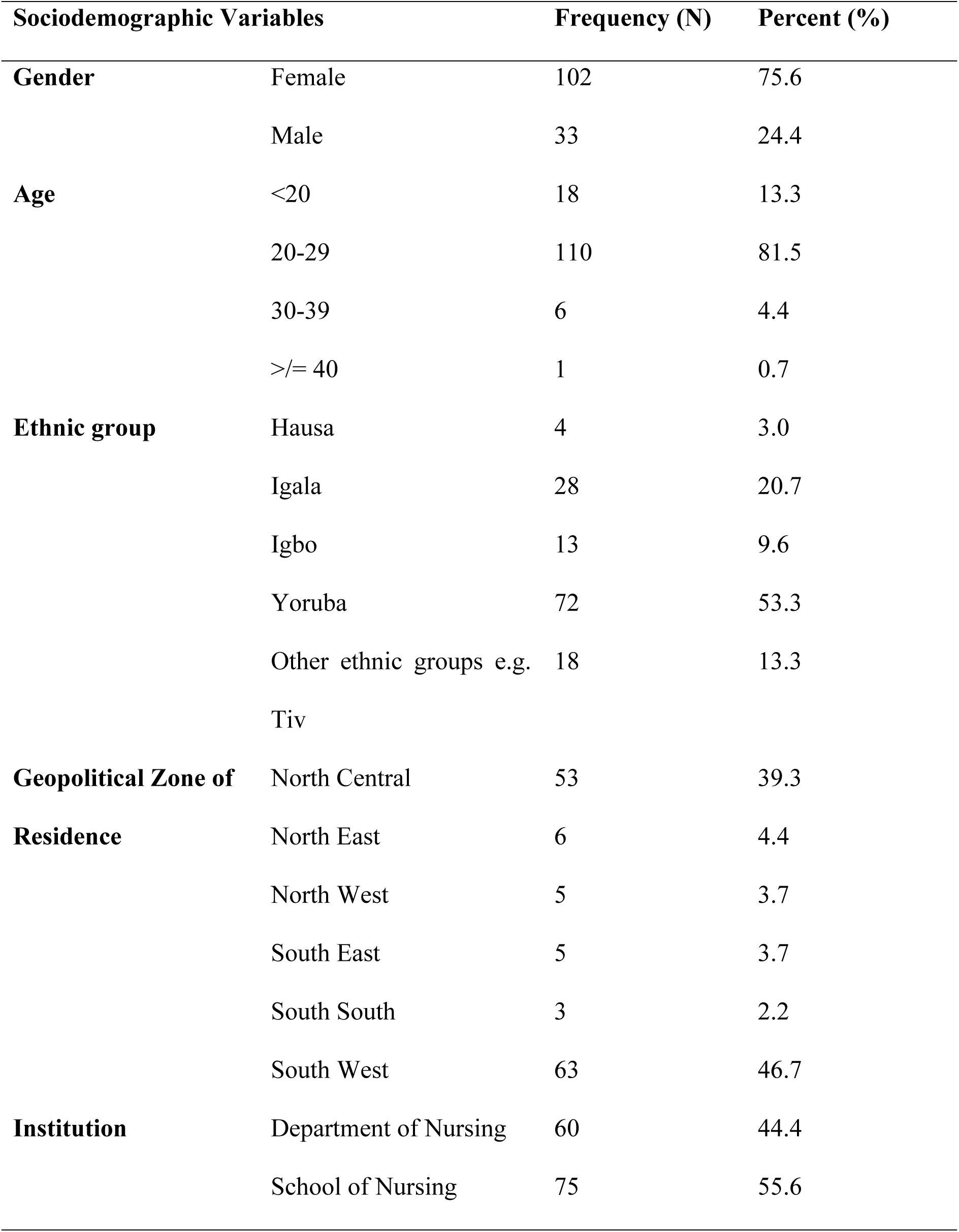
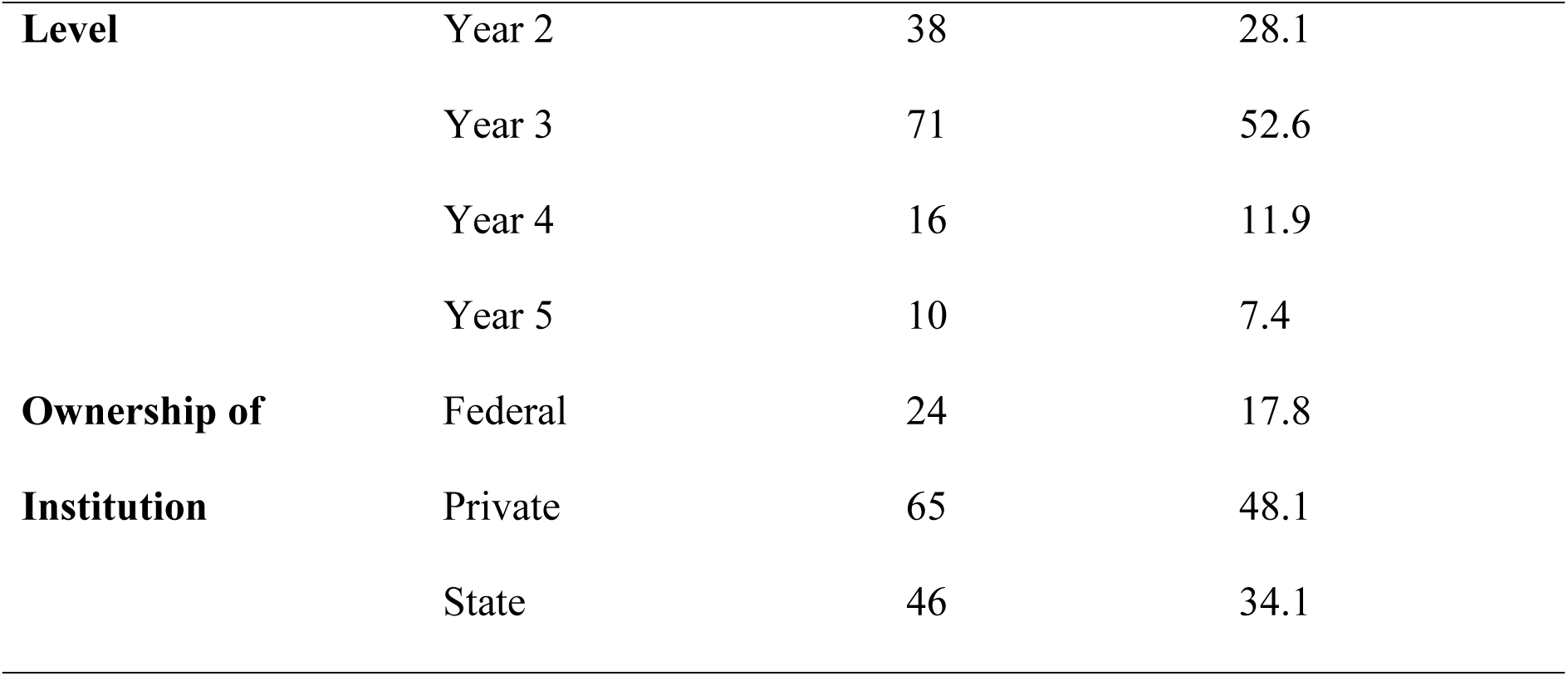
Sociodemographic information of the study population.

Table 2 shows the knowledge and perception of Nursing Students before and after online digital educational Intervention. The mean score of research knowledge before the intervention was 12.57 ± 2.90, while it increased to 14.21 ± 3.13 after the intervention. Similarly, research perception was 13.50 ± 3.38 before the intervention. However, this was slightly reduced after the intervention (12.75 ± 2.97).

**Table 2.** Outcome variables of the sample population before and after educational intervention.

| **Outcome Variables** | **Pre-test**<br>**(n=135)**<br>**Mean ± SD** | **Post-test**<br>**(n=135)**<br>**Mean ± SD** |
| --- | --- | --- |
| **Research Knowledge** | $12.57 \pm 2.90$ | $14.21 \pm 3.13$ |
| **Research Perception** | $13.50 \pm 3.38$ | $12.75 \pm 2.97$ |

Table 3 shows the categorization of research knowledge and perception before and after the educational intervention. The answers to the 25 questions of the knowledge scale were added to determine the knowledge scale scores, with a minimum of 0 and a maximum of 25 possible. Based on the scale, the population had an improved high level of research knowledge before and after the educational intervention (54 (40.0%) vs 83 (61.5%)). Similarly, the number of participants with a good perception level also increased after the educational intervention (128 (94.8%) vs 133 (98.5%).

**Table 3.** Categorization of research knowledge and perception before and after the educational intervention.

| **Variables** | **Level** | **Score** | **Pre-Test**<br>**n (%)** | **Post-Test**<br>**n (%)** |
| --- | --- | --- | --- | --- |
| Research Knowledge | Low | 0-13 | 81 (60.0%) | 52 (38.5%) |
|  | High | 14-25 | 54 (40.0%) | 83 (61.5%) |
| Research Perception | Poor | 0-7 | 7 (5.2%) | 2 (1.5%) |
|  | Good | 8-20 | 128 (94.8%) | 133 (98.5%) |

### Hypotheses testing

The knowledge and perception scores of nursing students in Nigeria towards research before and after an educational intervention were compared using the Mann-Whitney U test. See table 4

**Table 4:** Comparison of the scores of the outcome variables of the study population before and after the educational intervention.

| **Outcome Variables** | **Pre-test**<br>**(n=135)**<br>**Median (IQR<sup>a</sup>)** | **Post-test**<br>**(n=135)**<br>**Median (IQR)** | ***P*** | ***D<sup>b</sup>*** |
| --- | --- | --- | --- | --- |
| **Research Knowledge** | 13.00 (4.00) | 14.00 (5.00) | >.001 | 0.55 |
| **Research Perception** | 14.00 (5.00) | 13.00 (5.00) | .012 | 0.24 |

<sup>a</sup>Inter-quartile range
<sup>b</sup>Cohen's *d* effect size.

**H1:** There is a significant difference between the research knowledge of Nigerian nursing students before and after the intervention. The test showed a significant difference between the research knowledge of Nigerian nursing students before and after the intervention (p = <.001), with a medium effect size (*d* = 0.55). Hence, H1 was supported, which suggests that the post-test knowledge was significantly higher than the pre-test score.

**H2:** There is a significant difference between Nigerian nursing students’ perception of research before and after the intervention. The test revealed a significant difference between the research perception of Nigerian nursing students before and after the intervention *p* = .012, with a medium effect size of *d* = 0.24. Hence, H2 was accepted.

## Discussion

This study evaluated how an online digital educational intervention affected Nigerian nursing students’ knowledge and perception of research. According to Connor et al. [21], sound clinical judgment and nursing advancement are achieved through evidence obtained from research. Though Nigerian nursing students’ knowledge of research was lower before the intervention, a rise in the knowledge was recorded after the online digital educational intervention, consistent with the conclusions from research carried out by El-Khadry et al. [22], which shows a significant improvement in knowledge of research post-educational intervention. According to a review by Doran and Van de Mortel [34], digital learning activities were viewed favorably by students because they found the activities to enhance their learning. Also, in an experimental study by Ching et al. [23], learning achievement and learning satisfaction were found to be statistically significantly greater in the experimental group that used the online-based strategy than in the control group in the study.

The low level of knowledge of Nigerian nursing students before the intervention might be related to barriers such as a lack of proper education/training in the research process while in school, due to the complexity of the research curriculum, lack of previous studies, and further participation in research, lack of skilled supervision, etcetera. This conforms to the study by Akingbade et al. [9], which regarded these factors as the bedrock for a faulty foundation in research. Since the online digital educational intervention increased the participants’ knowledge, a more strategic plan should be implemented for a modified virtual learning platform to improve and simplify the research curriculum and introduce it during the early years of undergraduate levels. This will give students ample time to acquire adequate skills and research capacity [24,10]. A previous study [19], which evaluated the effectiveness of virtual classroom training in enhancing the knowledge and skills of general nursing and midwifery students, indicated that virtual classroom training significantly improved the knowledge and skills of this group, other studies have supported the notion that digital learning contributes to skill enhancement and overall learning improvement [25, 26].

In general, there is an increase in the level of perception towards research after the online digital educational intervention compared to the one recorded before the intervention. The participants perceived scholarly research as essential for evidence-based practice (EBP). This affirms the result obtained from research by Black et al. [27] and Keib et al. [28], who indicated that nursing students can have a positive change in perception towards research when they pass through the rudiments of being grounded in research courses and when taught from the preliminary stage of their baccalaureate program. Likewise, this is related to the study by Karim and Zeb [29], where most nursing students generally had a favourable opinion of the research and stated that it was beneficial to their field. This increase is significant as it reveals the importance of such educational intervention to help improve some nursing students’ research perception. However, Aderibigbe and Gbadamosi [30] posited that research is difficult and complex, which makes students’ participation in it minimal. This study revealed otherwise, as some participants neither considered research a difficult or boring field of study nor a time-waster; rather, a significant number of them thought it was just a part of the academic qualification needed as a nurse, which makes it a routine subject. This finding is supported by one of the studies conducted by Akingbade et al. [9], which reported that the participants also consider research as part of the minimum criteria to obtain the professional qualification to be a nurse, though they consider conducting research a stressful endeavour. The study showed a significant difference in how Nigerian nursing students perceive research before and after the educational intervention. Even though there are also little/no resources to support this evidence, it is believed that appropriate knowledge dissemination of the research process will further influence how they perceive research positively, which will determine the approach to conducting one. The result of this present study indicates that, with appropriate/simplified learning guides to research courses, mentorship [31], and research utilization, many Nigerian nursing students will see the need to improve their attitude toward acquiring more knowledge of research, hence building their confidence and enabling them to see research as rigorous, not difficult.

The study revealed a significant difference in the knowledge and perception of Nigerian nursing students about research when comparing the pre-and post-educational interventions. Overall, there is a limited study available to compare the research knowledge of nursing students before and after the intervention; meanwhile, when comparing the pre and post-educational interventions in some studies in other subject matters not related to nursing research, it was discovered that knowledge statistics significantly increased, a significant indicator that illustrates the fact that education improves learning (32-36]. This stance is also similar to a study by Doran and Van de Mortel [25], where the usefulness of an educational intervention in raising awareness, perception change, and boosting knowledge was demonstrated. Since the educational intervention of this current study was delivered online, and evidence suggests that there is a high usage of digital devices in Nigeria [37, 38], such format can be utilized as an adjunct to the conventional face-to-face mode of learning because, in online platforms, some students might be able to express their thoughts and ideas better than in a physical class [39]. Adesuyi et al. [26] suggested that nursing educational institutions can adopt online platforms to support the physical mode of learning, and this can improve learner-instructor interaction, which in turn aids better comprehension. However, it is important to address technical and internet issues that students might experience while using digital platforms [40]. On the contrary, as promising as online learning platforms are, Phirangee and Hewitt [41] suggested in their study that students should use non-verbal cues for effective communication.

With appropriate and guided research course content, an online research learning platform, and a reformed nursing educational system – that does not portray research as just an academic routine to credentialing but as a major part of the nursing curriculum – Nigerian nursing students may have an improved understanding of the research process and acquire the technical know-how to conduct credible research to improve nursing practice and for publication. This is consistent with the findings by Akingbade et al. [9], in which continuing education in research is emphasized.

Future studies could explore extended or repeated sessions to assess the long-term impact of similar educational interventions, which can yield more sustained knowledge retention and behavioural change. Likewise, the implications of this study’s findings are restricted to the Nigerian nursing education system and cannot be generalized to other environments or nations.

## Conclusion

This study demonstrates a notable rise in the knowledge of Nigerian Nursing students towards research post-educational intervention. This shows that the low level of knowledge before the online digital educational intervention can be traced to factors such as improper training and the complexity of the research curriculum. A good level of perception of nursing students in Nigeria towards research was established post-educational intervention in this study. This shows that there can be an improvement in the understanding, practice, and utilization of research in nursing practice in Nigeria when Nursing students are trained and well-equipped with adequate research knowledge. This will further improve the attitude of Nigerian Nursing students towards research and make them open to acquiring more knowledge and skills in research. A digital student-friendly approach can be developed to train nursing students in research to aid effective learning and practice, provide the best patient care, and achieve better outcomes. Although the knowledge and perception of nursing students were improved after the intervention, it is worthy of note to mention that the knowledge trajectory will diminish if interventions like this is not continuously put into actual actions/engagement. Likewise, the implications of this study’s findings are restricted to the Nigerian nursing education system and cannot be generalized to other environments or nations.

### Recommendation/Implication for Policy and Nursing Education

A good foundation of nursing research can be laid for nursing students right from the beginning of the baccalaureate program. Nursing students can be made to see that research is not just a part of the requirement for their academic qualification, but a major skill needed to practice effectively and efficiently as a nurse. The teaching of the research curricula can be simplified and considered to aid the students’ understanding. Academic mentors can be made available for students to guide and supervise the students until they master the skills in research. Likewise, students can be made to participate in research conferences where they can relate with other researchers and have the research rudiments explained thoroughly. Lecturers and supervisors can be trained and equipped with the necessary skills to help the students advance and communicate their needs.

### Strengths and limitations of this study

We acknowledge that conducting the study with a one-day online educational intervention may introduce certain limitations to the study’s validity, particularly in terms of the depth and retention of knowledge gained over such a short period. However, the intervention was designed to be concise and focused to maximize engagement and minimize participant burden, which can be particularly important in online formats. To address potential validity threats, we incorporated pre- and post-intervention assessments to measure immediate learning outcomes and comprehension. Additionally, we plan to discuss these limitations in our study findings, noting that a longer intervention might yield more sustained knowledge retention and behavioural change. Future studies could explore extended or repeated sessions to further assess the long-term impact of similar educational interventions. In addition, since the data was collected online, coupled with internet fluctuations, connecting with people physically or directly is impossible, limiting participants’ response rate.

## Data Availability

The dataset used and analyzed supporting this study findings are available upon reasonable request from the corresponding author.

## Acknowledgments

This study acknowledges all the nursing students who participated in the webinar and research for their support.

## Notes

### Competing Interest Statement

The authors have declared no competing interest.

### Funding Statement

The author(s) received no specific funding for this work.

### Author Declarations

The Lagos University Teaching Hospital Health Research Ethics Committee, Idi-Araba, Lagos State, granted ethical approval to conduct this study (ethical approval number ADM/DSCST/HREC/APP/6322)

